# Exploratory dried blood spot metabolomics identifies pathway-level convergence with ME/CFS biology in a self-reported PEM-like fatigue phenotype

**DOI:** 10.64898/2026.06.08.26355197

**Authors:** Pierrick Hauguel, Nicolas Anctil, Louis-Philippe Noel

## Abstract

**Background:** Plasma and serum metabolomic studies of myalgic encephalomyelitis / chronic fatigue syndrome (ME/CFS) have repeatedly implicated hypometabolic, lipid, mitochondrial, redox and tryptophan-kynurenine pathways, but prior cohorts have been modest in size and have used heterogeneous case definitions. Whether similar pathway-level signals are detectable at scale in dried blood spots (DBS), across questionnaire-derived fatigue constructs and across orthogonal LC gradients in the same individuals remains unresolved.

**Methods:** We profiled DBS extracts from 1,784 community-cohort adults by reverse-phase LC-MS using paired 5 min and 15 min gradients. Six questionnaire-derived endpoints captured a pragmatic self-reported PEM-like phenotype, a DSQ-derived PEM-like construct, high or review clinical status, temporal fatigue state, comorbid fatigue and self-reported chronic fatigue. The locked primary endpoint for Phase 1 was `pragmatic_fatigue_pem` with 226 cases and 914 controls after excluding major metabolic comorbidity. We tested a biology-first panel comprising 22 literature-curated metabolites represented by four participant-level descriptors each, and evaluated three discovery extensions: a targeted m/z search of additional literature candidates, a hypothesis-free univariate screen across 4,553 5 min and 5,625 15 min consensus features, and pairwise z-difference ratios. Endpoint-specific Ridge classifiers were evaluated by five-fold out-of-fold AUC with bootstrap stability filtering. Cross-gradient agreement was assessed by per-metabolite AUC concordance between paired 5 min and 15 min profiles. Severity was modelled as an ordinal grade derived from the number of fatigue criteria met and chronic-fatigue-form status.

**Results:** The biology-first DBS panel achieved out-of-fold AUC 0.81 for the pragmatic self-reported PEM-like endpoint (226 cases / 914 controls). The DSQ-derived PEM-like construct reached AUC 0.60 (57 cases / 201 controls) on the un-filtered set and AUC 0.778 (SD 0.013, twenty seeds) in a post-hoc signature-decomposition follow-up restricted to participants without a self-declared major-metabolic-history tag (29 cases / 230 controls); both are treated as construct-validity anchors rather than as provoked or clinically adjudicated PEM. An optimised operationalisation of the same construct (panel-self normalisation, restriction to non-comorbid participants and demographic covariates) reached AUC 0.71 (95 % CI 0.55 to 0.76), and an exploratory age-stratified signature decomposition suggested age-dependent pathway composition that requires confirmation given small per-stratum case counts. Stable contributors mapped to carnitine-shuttle, TCA-cycle, redox-thiol and tryptophan-kynurenine pathways. Cross-gradient analysis of 22 matched metabolites yielded Pearson r = 0.62 for signed univariate effects (p = 0.002; 68 % directional agreement). The metabolomic score increased with severity grade (Spearman rho = 0.45, p = 4 × 10^−91^; median scores 0.24, 0.51 and 0.75 across grades 0, 1 and 2). Sensitivity analyses on the covariate-complete subset (n = 565; 138 cases / 427 controls) showed that the DBS signal was robust to adjustment for age, sex, BMI and medication burden (DBS-only AUC 0.76, DBS plus covariates 0.78, covariates only 0.64), and produced a metabolomic-specific lift of approximately 0.13 AUC over the strongest anti-leak declarative cross-form questionnaire baseline (AUC 0.63). DBS-only AUC was stable across sex, age and BMI subgroups, and a 1:4 nearest-neighbour matched analysis on age, sex and BMI yielded AUC 0.72 (95 % CI 0.67 to 0.77). The observed pattern supported pathway-level convergence with prior ME/CFS metabolomics literature, including carnitine shuttle, fatty-acid beta-oxidation, TCA cycle, redox-thiol, urea cycle, glycerophospholipid and tryptophan-kynurenine axes. In contrast, the hypothesis-free 15 min screen produced high-AUC features that mapped predominantly to environmental or technical signals, including pesticide, industrial-amine and mobile-phase artifact annotations; only one of eight top leads, a truncated oxidised phospholipid, was biologically plausible, and none had tandem-MS support.

**Conclusions:** In this large community cohort, a literature-curated DBS metabolomic panel captured pathway-level biology associated with a questionnaire-derived PEM-like fatigue phenotype, showed directional concordance across LC gradients, scaled with symptom severity and remained robust to key demographic, anthropometric and anti-leak questionnaire baselines. The findings converge with several metabolic axes previously reported in ME/CFS plasma and serum studies, including carnitine-shuttle, TCA-cycle, redox-thiol, urea-cycle, glycerophospholipid and tryptophan-kynurenine pathways. They should not be interpreted as clinical validation of a diagnostic test, screening tool or objective provoked-PEM biomarker. Rather, they support at-home-compatible DBS metabolomics as a biologically grounded platform for future clinically adjudicated validation, decision-support development and longitudinal monitoring in fatigue and PEM-like syndromes. Because DBS contains cellular and plasma-derived components, matrix effects must be considered when comparing individual metabolites with venous plasma or serum studies, and hypothesis-free screening at this scale can preferentially surface exposome or technical variance unless molecular identification is enforced before biological interpretation.

## 1 Introduction

Myalgic encephalomyelitis / chronic fatigue syndrome (ME/CFS) is a multi-system illness whose cardinal feature is post-exertional malaise (PEM), defined by the 2015 Institute of Medicine criteria as a disproportionate worsening of symptoms after physical, cognitive or emotional exertion that may persist for 24 hours or more [1]. The DePaul Symptom Questionnaire (DSQ), originally developed to operationalise the Canadian Consensus Criteria, remains one of the most widely used self-report instruments for research case finding and symptom grading [2]. Despite the disease burden, no molecular biomarker has been validated for diagnostic use. Clinical heterogeneity, comorbidities and inconsistent case definitions have also complicated mechanistic studies, as illustrated by a systematic review identifying 20 published case definitions for CFS/ME [3, 4].

A series of plasma and serum metabolomic studies has nonetheless converged on a pathway-level hypometabolic pattern. Early work reported acylcarnitine deficiencies in CFS patients, implicating impaired fatty-acid transport into mitochondria [5, 6], and broader metabolomic profiling later highlighted anomalous energy metabolism and oxidative-stress pathways [7]. Naviaux and colleagues reported a multi-pathway plasma signature in which most discriminatory metabolites were lower in cases, with strongest signals in sphingolipids, phospholipids, glycosphingolipids, branched-chain amino acids and mitochondrial metabolism [8]. Yamano et al. identified ornithine/citrulline and pyruvate/isocitrate ratios as discriminators, implicating TCA and urea-cycle dysfunction [9]. Germain and colleagues reported fatty-acid beta-oxidation, glycerophospholipid and acyl-lipid disruption [10, 11], a redox-thiol imbalance signature [12], and a distinctive 24-hour plasma response to maximal exercise [13]. Nagy-Szakal et al. integrated metabolomic and faecal metagenomic data into a combined predictor highlighting choline, carnitine and ceramide axes [14]. Che et al. reported decreased plasmalogens, phosphatidylcholines and sphingomyelins together with elevated dicarboxylic acids, providing metabolomic evidence for peroxisomal dysfunction in ME/CFS [15]. Across these reports, the tryptophan-kynurenine pathway has emerged as a candidate unifying axis [16], with related signals in long-COVID studies that have renewed interest in ME/CFS-like post-acute syndromes [17-20].

However, methodological variability has prevented convergence on a consensus metabolite list and no published plasma cohort has exceeded 200 cases [4]. Large-scale DBS assessment across alternative fatigue definitions and orthogonal LC gradients in the same individuals has not been reported.

Here, we analysed a community cohort of 1,784 adults profiled from fingerstick DBS extracts in paired 5 min and 15 min reverse-phase LC-MS gradients. The DBS format is compatible with home self-collection by fingerstick without phlebotomy, which is operationally relevant in fatigue and PEM-like syndromes where repeated clinic attendance can be burdensome or inaccessible for the most affected individuals. We aimed to quantify the performance of a biology-first DBS panel curated from the prior ME/CFS metabolomics literature across six fatigue endpoints, test whether pathway-level signals reported in plasma and serum cohorts are detectable in a cellular-rich capillary blood matrix, assess technical concordance through cross-gradient analysis, evaluate whether the metabolomic score scales with symptom severity, and examine the interpretability of hypothesis-free univariate screening when feature identification is performed before biological interpretation.

## 2 Methods

### 2.1 Cohort and ethics

The study included 1,784 adult participants enrolled through the BioTwin program. Participants provided written informed consent for fingerstick blood collection and structured health questionnaires. Samples were collected on dried blood-spot cards and stored frozen before LC-MS analysis. No plasma-separation device was used; the analysed matrix is therefore DBS rather than venous plasma. Fingerstick DBS sampling is compatible with home self-collection without phlebotomy, and a subset of participants performed repeated self-collected sampling between in-person visits. The study was approved by the Canadian SHIELD Ethics Review Board (REB Tracking Number: 2023-11-003; OHRP Registration IORG0003491; FDA Registration IRB00004157; initial approval granted December 15, 2020; continuing review approval granted April 30, 2025, valid through April 29, 2026). Ethical reporting follows the Declaration of Helsinki [21].

### 2.2 LC-MS profiling

DBS extracts were profiled on the same instrument using two reverse-phase LC gradients: a 5 min production gradient used for routine reporting and a 15 min deep gradient used to improve low-abundance and isobaric resolution. Detection was performed in positive ion mode. Peak picking, feature alignment and quantification were performed with OpenMS [22]. After coverage filtering, the 5 min run yielded 4,553 consensus features and the 15 min run yielded 5,625 consensus features.

### 2.3 Feature engineering

Per-card feature intensities were log1p-transformed and converted to robust z-scores within each feature-by-batch stratum using median centering and median absolute deviation scaling, with a standard-deviation fallback for low-variance strata. Participant-level descriptors were computed by aggregating per-card z-scores within each participant-feature group as the median, 90th percentile, inter-quartile range and number of non-missing cards. The biology-first panel comprised 22 literature-curated metabolites represented by four engineered descriptors each, yielding 88 participant-level model inputs. These descriptors should be interpreted as repeated summaries of 22 metabolites, not as 88 independent biochemical analytes.

### 2.4 Endpoint definitions

Six endpoints were derived from questionnaire data. The pragmatic endpoint used a three-criterion operationalisation of substantial activity reduction, post-exertional symptom worsening and unrefreshing fatigue, approximating the IOM 2015 cardinal triad [1]. This endpoint was selected as the powered PEM-like construct, whereas the DSQ-derived PEM-like construct provided a smaller construct-validity anchor. These definitions are not interchangeable with clinically adjudicated ME/CFS or provoked PEM.

- **Pragmatic PEM-like phenotype:** 2 of 3 pragmatic pillars (226 / 914, exclude major metabolic comorbidities). Strict controls: 0 pillars, no chronic fatigue form, no metabolic comorbidity, not v1 hardcoded CFS.
- **DSQ-derived PEM-like construct:** DSQ-based PEM-like positive (57 / 201) [2].
- **Temporal fatigue state:** high temporal fatigue label (153 / 105).
- **High/review clinical status:** high or review status from internal review flags (129 / 129).
- **Comorbid fatigue:** fatigue with comorbid metabolic context (60 / 1,007, strict controls).
- **Chronic fatigue form:** self-declared chronic fatigue diagnostic form (12 / 246, included for completeness; n_pos too small for confident multivariate inference).

### 2.5 Questionnaire anti-leak handling

Questionnaire variables were separated into two layers before interpretation. DSQ, chronic-fatigue and SF36 fatigue variables define or cross-check a self-reported PEM-like phenotype. Daily, morning, daytime, evening and sample-card forms provide temporal state and sample context. They should not be mixed as clinical validation.

The Phase 1 anti-leak matrix was resolved on 2026-05-26 (`phase1_outputs/phase1_a2_questionnaire_items_` Direct PEM, fatigue, sleep, chronicity, clinical-review and temporal-state variables are excluded from predictive baselines for endpoints they helped define. The next modeling step should analyse two separated questionnaire baselines: a clean covariate baseline using demographics, anthropometrics, smoking, alcohol and derived medication burden; and a declarative cross-form baseline using only allowed cross-form symptom variables by endpoint. The cross-form baseline can assess self-report concordance, but it is not an independent clinical validation.

### 2.6 Discovery extensions

**Wave A:** a literature-targeted m/z search of 80 candidate metabolites returned 120 retained matches, mapped to 106 unique consensus features (48 in 5 min, 72 in 15 min) across 23 pathway labels, including carnitine shuttle, TCA cycle, BCAA, redox-thiol, tryptophan-kynurenine, sphingolipid, stress-steroid and polyamine pathways.

**Wave B:** a hypothesis-free univariate rank-AUC screen tested all consensus features with adequate coverage (4,553 in 5 min, 5,625 in 15 min) against the six endpoints. Features were ranked by their absolute centred AUC across endpoints and required cross-stability, defined as top-k overlap ≥ 50 % across 60 bootstrap resamples. Twenty-three 5 min features and four 15 min features met the cross-stability threshold at AUC ≥ 0.60 on all six endpoints.

**Wave C:** pairwise z-difference ratios were generated from the top 80 features (3,160 ratios) and screened in the same Ridge framework. Nine ratios met bootstrap stability criteria on at least two endpoints; the multivariate gain over the feature pool alone was marginal.

### 2.7 Models and evaluation

Endpoint-specific Ridge classifiers (logistic regression with L2 penalty, balanced class weights, C approximately 0.1) were trained with five-fold stratified cross-validation using the scikit-learn implementation [23]. Missing feature values were imputed as zero after robust z-scoring. Out-of-fold AUC and average precision were the primary performance metrics. Bootstrap stability filtering (60 to 100 iterations, top-k feature selection, selection-frequency threshold 0.40 to 0.50) was used to identify stable contributors to the biology-first and Wave A panels. All AUC values reported in the paper are out-of-fold unless otherwise stated.

### 2.8 Cross-gradient analysis

For each of the 22 biology-first metabolites identified in the 5 min gradient, the 15 min consensus feature table was searched for matches within ± 5 ppm m/z. Matches seen in at least 200 samples were retained to limit the analysis to adequately covered features. The best-covered 15 min match for each metabolite was carried forward. Participant-level z-scores were rebuilt independently for each gradient, and per-metabolite univariate AUC against the pragmatic PEM-like endpoint was computed in both gradients. Concordance was reported as the Pearson correlation of AUC – 0.5 across the 22 metabolites and as the proportion of metabolites with the same directional effect in both gradients.

### 2.9 Severity stratification

Severity was encoded as an ordinal grade: 0 (strict control: 0 pragmatic pillars, no chronic fatigue form, no major metabolic comorbidity); 1 (1 pragmatic pillar); 2 (2 pillars, pragmatic case); 3 (3 pillars); and 4 (chronic fatigue form). For participants in the locked Phase 1 case-control set (226 cases and 914 controls), the out-of-fold biology-first model probability was used as the metabolomic score. For remaining participants, the probability from a model trained on the full case-control set was projected for visualisation. Trend was tested with Spearman correlation and group differences with Kruskal-Wallis and Mann-Whitney U tests.

### 2.10 Identification of Wave B 15 min leaders

The four 15 min Wave B leaders that satisfied cross-stability at AUC ≥ 0.60 on all six endpoints, plus four follow-up candidates from the next ranks, were searched against the platform identification table by exact m/z match. Tandem-MS evidence was queried in three independent MS2 result directories generated by the o1.5 pipeline (`ms2_identification`, `ms2_identification_v2`, `ms2_propagation`) using tolerances up to ± 10 ppm m/z and ± 30 sec retention time. None of the eight leaders had MS2 support in any directory. Identification confidence is therefore reported following the Schymanski et al. framework [24]: these leads remain at Level 4 or Level 5 until tandem-MS spectral matches are obtained.

### 2.11 Sensitivity analyses and anti-leak questionnaire baselines

Sensitivity analyses were run on the locked primary endpoint (`pragmatic_fatigue_pem`; 226 cases, 914 controls), using the same model spec as the primary panel to keep AUCs directly comparable: `Simpleimputer(median)` then `RobustScaler` then `RidgeClassifier(class_weight=“balanced”, alpha=10.0)` evaluated by five-fold stratified out-of-fold cross-validation, with AUC computed on `sigmoid(decision_function)`. Bootstrap 95 % CIs were obtained from 1,000 resamples of the out-of-fold predictions.

Seven panels were evaluated, all on the same model spec. (A) DBS-only on the full eligible set (88 biology-first descriptors, n = 1,140). (B) DBS-only restricted to participants with plausible-range covariates (`covariate_complete_core_qc`, n = 565; 138 cases / 427 controls), to allow direct comparison with covariate-adjusted models. (C) DBS plus four covariates (age, sex, BMI, log1p of an additive medication burden) on the QC subset. (D) Covariates only, on the QC subset, as a demographic ceiling. (E) An anti-leak clean covariate baseline (age, sex, BMI, BMI^2, smoking, weekly alcohol drinks, medication burden) using only items classified `allow_primary_covariate` for this endpoint in the 2026-05-26 anti-leak matrix. (F) An anti-leak declarative cross-form baseline (E plus the two cross-form items that pass the anti-leak filter for this endpoint: `dsq2_cog_n_endorsed`, allow_except_dsq_clinical_review; `sf36_general_health_severity_max`, allow_except_review). (G) F plus four `allow_sensitivity_only` items (`dsq1_onset`, `dsq1_diagnosed`, `cf_duration_n`, `cf_has_clinical_anchor`) as an upper-bound questionnaire context.

Subgroup robustness was assessed by computing DBS-only out-of-fold AUC stratified by sex, age (<50 vs ≥ 50) and BMI (<25, 25 to 30, ≥ 30). A matched-control analysis nearest-neighbour matched each pragmatic PEM-like case to up to four controls without replacement on age (± 3 years), sex (exact) and BMI (± 2 kg/m^2); the DBS-only model was refit out-of-fold on the matched set.

## 3 Results

### 3.1 Cohort and pipeline (Figure 1)

Of 1,784 adults, the locked Phase 1 pragmatic PEM-like definition produced 226 cases and 914 strict controls after excluding major metabolic comorbidities; the remaining endpoints had between 12 and 153 cases each. The analysis workflow combined the biology-first DBS panel with three discovery extensions and out-of-fold Ridge evaluation across all six endpoints.

**Figure 1.**
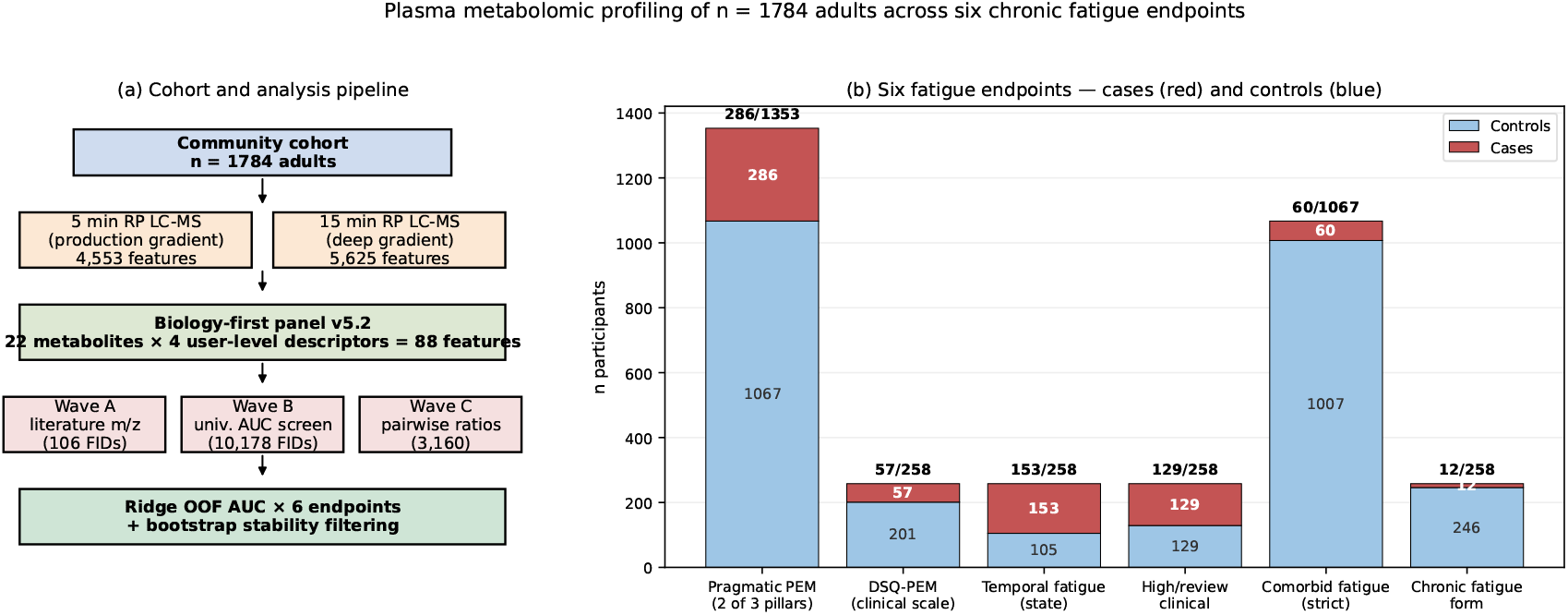
Cohort, analysis workflow and endpoint cardinalities.

### 3.2 Endpoint specificity (Figure 2)

The biology-first panel achieved an out-of-fold AUC of 0.81 on the pragmatic PEM-like end-point (226 / 914). Performance ranked: pragmatic PEM-like phenotype (0.81), comorbid fatigue (0.73), chronic fatigue form (0.71; 12 cases, see limitations), temporal fatigue state (0.65), high/review clinical status (0.60) and DSQ-derived PEM-like construct (0.60). This creates an explicit trade-off: the pragmatic endpoint provides statistical power, whereas the DSQ-derived construct provides stronger questionnaire construct anchoring but substantially lower case count. The best-performing endpoints encoded either explicit PEM-pillar logic or metabolic-comorbidity context. Generic temporal fatigue and high/review status carried less metabolic signal than PEM-like case definitions.

**Figure 2.**
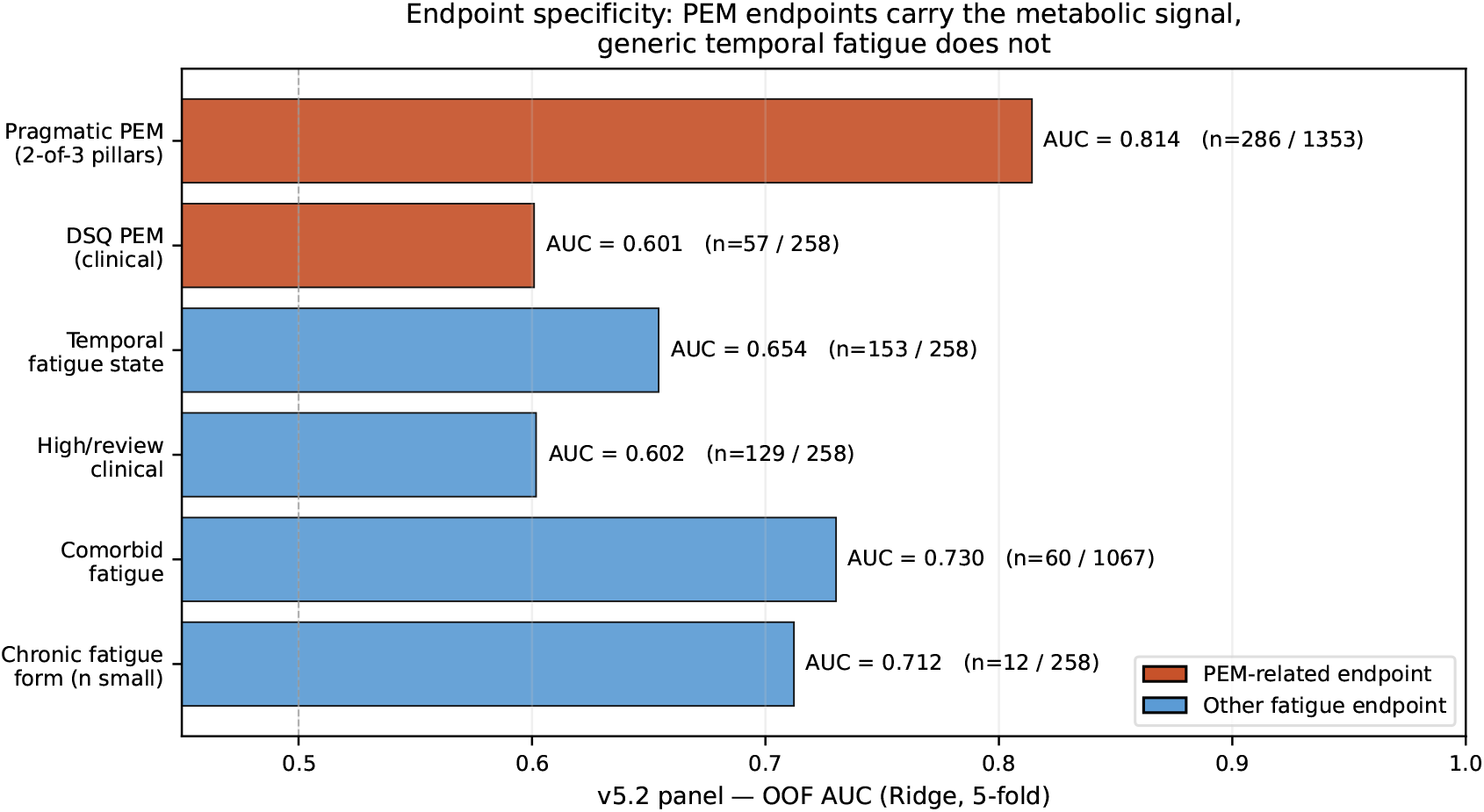
Out-of-fold AUC by fatigue endpoint for the locked DBS biology-first panel.

#### DSQ-PEM follow-up by signature decomposition

A post-hoc exploratory analysis decomposed the biology-first panel into four pathway-coherent signature blocks (S1 tryptophan-kynurenine, 7 features; S2 purine recovery, 3 features; S4 mitochondrial TCA and short-chain energy, 5 features; S5 glucogenic amino acids, 6 features), each modelled as four participant-level descriptors (median, 90th percentile, IQR and number of non-missing cards) plus age, sex and BMI. Out-of-fold predictions of the four per-signature Ridge models (`RidgeClassifier`, alpha = 10, class-balanced) were averaged across signatures (late fusion) on the DSQ-PEM-eligible set restricted to participants without a self-declared major-metabolic-history tag (`has_major_meta =` False; n - 259, 29 cases / 230 controls). Across twenty random seeds, late-fusion out-of-fold AUC reached 0.778 (SD 0.013), compared with 0.60 for the full 88-feature panel evaluated on the un-filtered 57 / 201 DSQ-PEM endpoint reported above. The improvement was robust to leave-one-out exclusion of the dense-sampling case (BT1018; AUC without BT1018 0.760, SD 0.014). On the same non-comorbid set (the “C6” cohort defined below), evaluation per-card with five-fold GroupKFold on participant identifier reached AUC 0.64, materially below the user-level estimate, indicating that the construct holds at the participant level (aggregated across multiple sampling occasions) and should not be deployed as a single-card readout. The `n_nonmissing_cards` descriptor contributed approximately 0.02 AUC in aggregate to the late-fusion model, but on the age < 40 sub-stratum univariate card-count AUC alone reached 0.86, so this descriptor is reported separately and is held out of the late-fusion estimate when interpreting age-stratified results. The signature decomposition is a post-hoc exploratory follow-up on a smaller cohort and is reported as a construct-validity anchor for the DSQ-PEM endpoint, not as a replacement for the headline pragmatic estimate.

#### DSQ-PEM operationalisation: an improved specification

Separately from the signature decomposition, the DSQ-PEM operationalisation itself was optimised relative to the locked specification. The locked model (full 88-descriptor panel, feature-by-batch robust z-scoring, full DSQ-positive versus screened-control contrast) reached mean out-of-fold AUC 0.588 (SD 0.023; twenty seeds; n = 315, 57 cases / 258 controls). Three independent, stackable adjustments each improved it: (i) per-sample normalisation computed against the 22-metabolite panel itself, combined with rank-z scaling within batch (mean AUC 0.638, SD 0.025; n = 299, 56 cases; -0.05 over locked); (ii) restricting both cases and controls to participants without a self-declared major-metabolic-history tag (mean AUC 0.651, SD 0.027; n = 287, 29 cases; -0.06 over locked); and (iii) adding age, sex and BMI as model covariates (+0.05 AUC within the non-comorbid set). Stacked together (panel-self normalisation, rank-z within batch, non-comorbid restriction and demographic covariates), this specification (labelled “C6”) reached mean AUC 0.708 (SD 0.027; bootstrap 95 % CI 0.55 to 0.76; n = 259, 29 cases / 230 controls), a 0.12 AUC gain over the locked specification on the same construct. The four-signature late fusion reported above (mean AUC 0.778) is this same non-comorbid cohort modelled by pathway block rather than as a single pooled classifier. Adjustments that did not help, and are not carried forward, included total-ion-current and pooled-external-feature normalisation, sample weighting by card count, and linear residualisation of features on age, sex and BMI (each neutral to -0.08 AUC). Per-variant results are in Supplementary S12. These remain post-hoc construct-validity analyses on a reduced non-comorbid cohort, not a clinically validated diagnostic threshold.

#### Exploratory age stratification suggests age-dependent PEM-like phenotypes

Refitting the per-signature late fusion within age strata of the non-comorbid set indicated that both the strength and the pathway composition of the DSQ-PEM separation varied with age. In adults aged 40 years and over (n = 181, 21 cases), signature decomposition recovered separation that the pooled panel did not: a stratum-specific late fusion of the mitochondrial-energy, tryptophan-kynurenine and glucogenic-amino-acid blocks reached mean AUC 0.717 (SD 0.027; CI 0.61 to 0.83), versus 0.623 for the full 88-descriptor panel in the same stratum. In participants under 40 (n = 49, 7 cases) a two-block fusion (gut-liver hippurate plus mitochondrial energy) reached mean AUC 0.94 (SD 0.02), and in participants 60 and over (n = 91, 7 cases) a glucogenic-amino-acid plus mitochondrial fusion reached mean AUC 0.68. The dominant block shifted from a broad multi-axis pattern in younger participants toward amino-acid and purine-recovery axes in older participants. Both extreme strata rest on only seven cases each, with correspondingly wide confidence intervals; these stratified results are exploratory, were not pre-specified, and are reported as hypothesis-generating rather than as stable phenotype estimates pending independent confirmation. Per-stratum, per-signature AUCs are in Supplementary S12.

### 3.3 Pathway-level findings (Figure 3)

Wave A pathway blocks added measurable lift on metabolic-burden endpoints, including comorbid fatigue (AUC 0.66 to 0.70), but did not improve over the biology-first panel on the pragmatic PEM-like endpoint. This suggests that the 22-metabolite curated panel had already captured most of the available signal for this endpoint. Carnitine-shuttle, TCA-cycle, redox-thiol and tryptophan-kynurenine blocks were the most consistent contributors across endpoints (Figure 3; AUC range 0.50 to 0.69 across 28 pathway-aware blocks).

**Figure 3.**
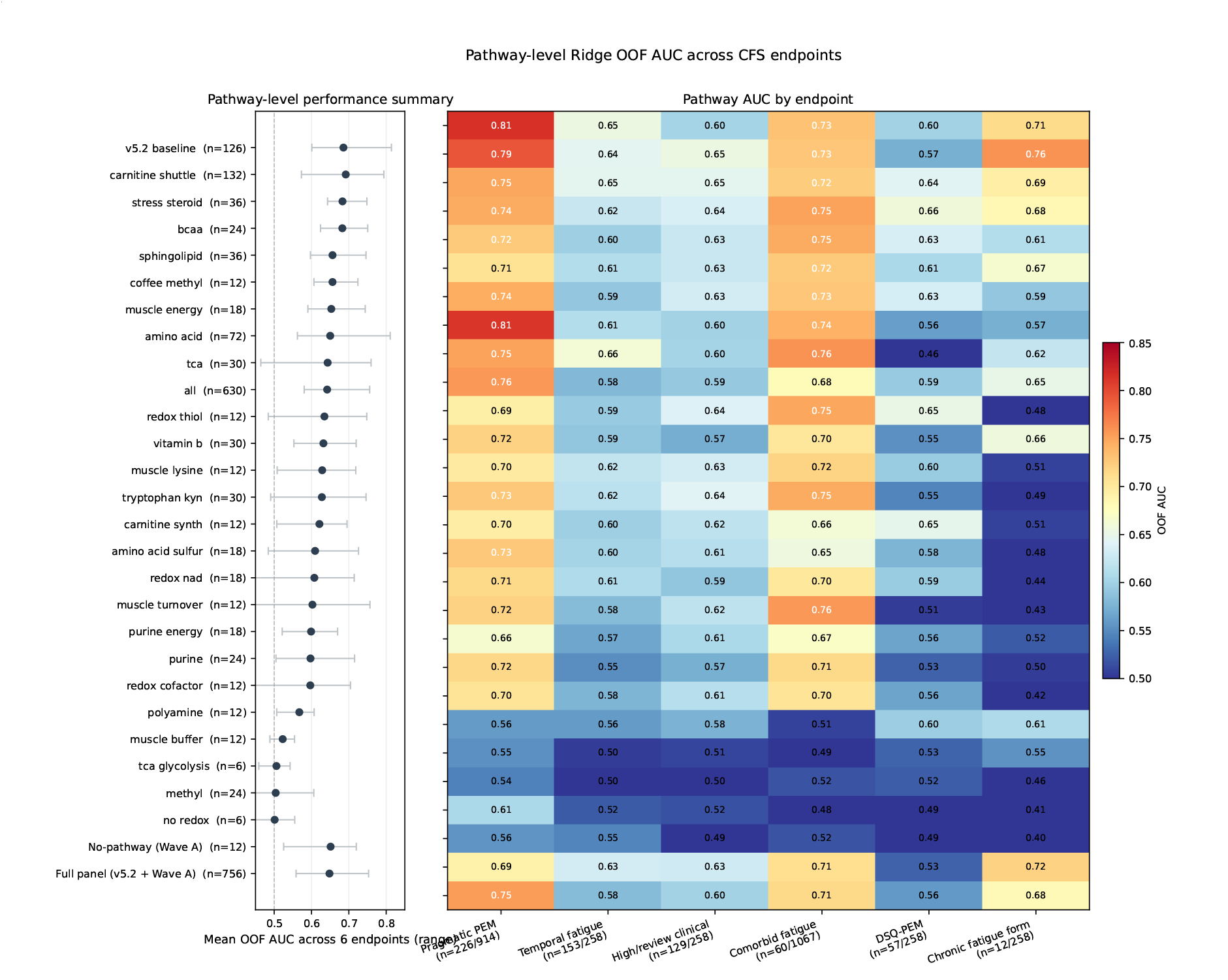
Pathway-level Wave A feature-block performance across fatigue endpoints.

### 3.4 Pathway convergence with prior cohorts (Figure 4b)

Six of the eight pathway axes that were significant in our DBS cohort had been previously reported with the same direction in at least one of Naviaux 2016 [8], Germain 2017 to 2020 [10, 11] or Che 2022 [15]: carnitine shuttle [5, 6], fatty-acid beta-oxidation [8, 10], TCA cycle [8, 9], redox-thiol [12, 15], urea cycle [9] and tryptophan-kynurenine [15, 16]. This is a pathway-level cross-matrix comparison rather than a claim of one-to-one metabolite equivalence with venous plasma or serum. Redox-thiol signals in DBS, including glutathione-related features, may include a substantial erythrocyte or cellular contribution. Sphingolipids and BCAA, the two strongest signals in Naviaux 2016 [8], were not significantly displaced in our cohort, consistent with both matrix and chemistry-coverage differences between targeted lipid panels, venous plasma/serum and a reverse-phase DBS small-molecule platform. Glycerophospholipids were depressed in our cohort and in Germain 2017/2020 [10, 11] and in the peroxisomal-dysfunction profile reported by Che 2022 [15].

**Figure 4.**
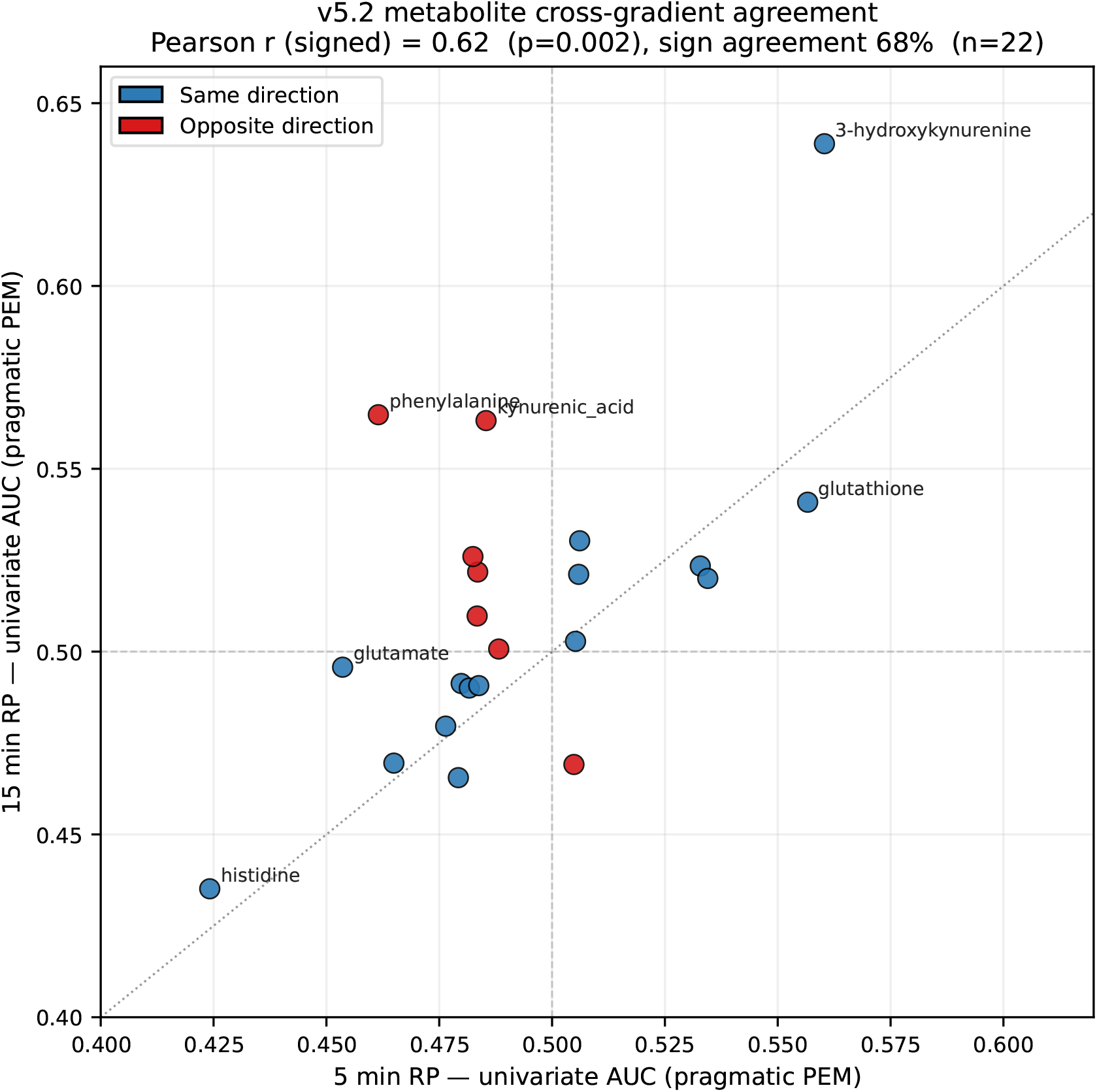
Cross-gradient agreement between 5 min and 15 min LC-MS gradients for 22 biology-first metabolites.

### 3.5 Cross-gradient agreement on the same individuals (Figure 4)

For the 22 biology-first metabolites, a 15 min counterpart was identified by m/z ± 5 ppm and per-metabolite univariate AUC on the pragmatic PEM-like endpoint was computed independently in each gradient. Signed effects were correlated at Pearson r = 0.62 (p = 0.002; approximate 95 % CI 0.27 to 0.83), with 68 % of metabolites showing the same direction (Figure 4). This provides technical concordance in the same participants: the same biological samples, acquired through two distinct chromatographic gradients, showed concordant directional effects for the panel metabolites. The strongest 5 min signals (histidine lower, 3-hydroxykynurenine higher, glutathione higher, glutamate lower) showed the same direction in the 15 min gradient; phenylalanine was the main exception, with a stronger positive signal in the 15 min gradient.

**Figure 4b.**
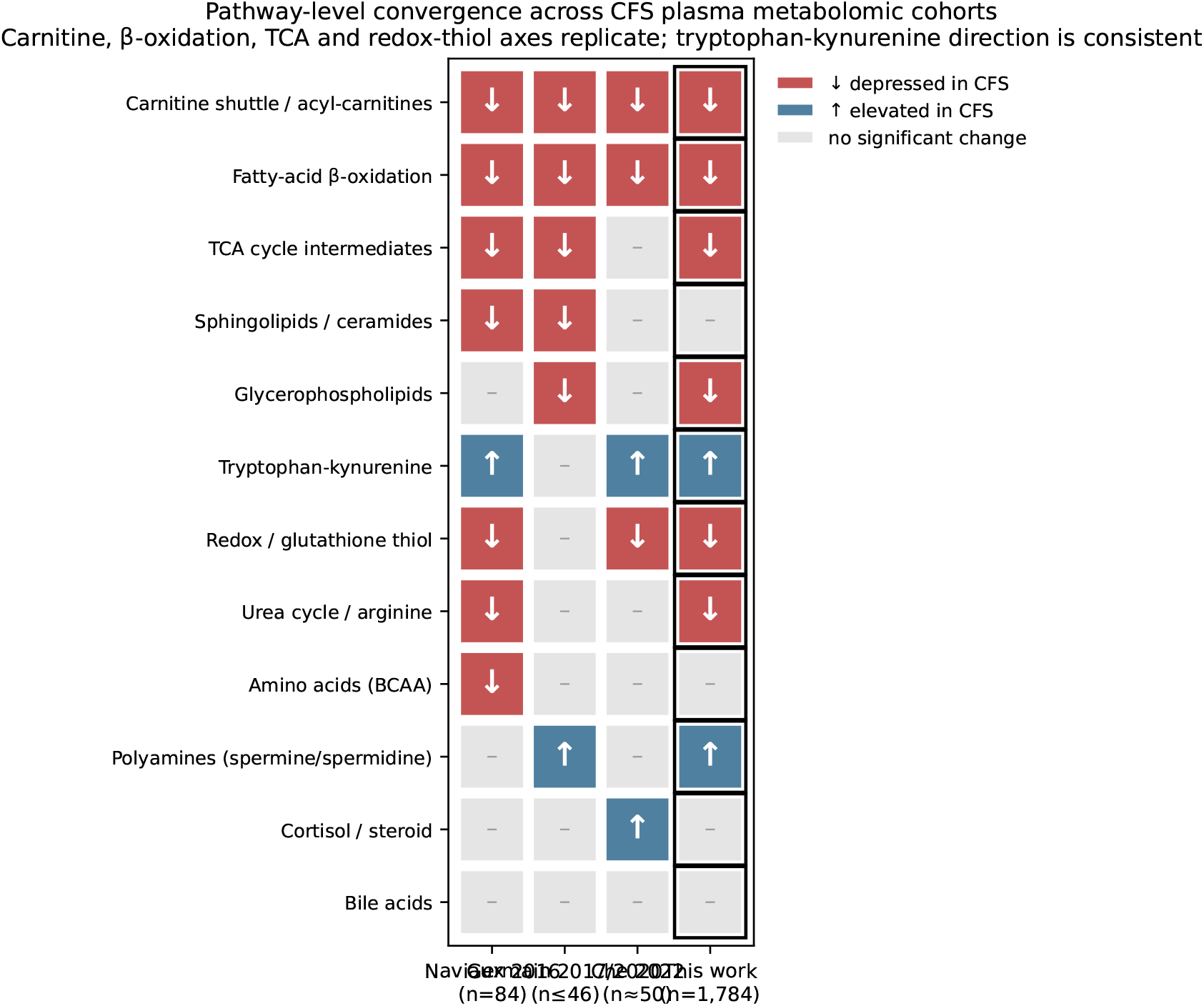
Pathway-level convergence between BioTwin DBS findings and prior ME/CFS metabolomics literature.

### 3.6 Severity stratification (Figure 5)

The biology-first score increased monotonically with ordinal severity grade across the full cohort (Figure 5; medians 0.24, 0.51 and 0.75 for grades 0, 1 and 2; Spearman rho = 0.45, p = 4 × 10^−91^; Kruskal-Wallis H = 378). Each step away from strict controls produced a significant median increase (Mann-Whitney p ≤ 5 × 10^−4^ for all grade-vs-control contrasts), including the small grade 4 chronic-fatigue-form group (n = 15, delta median = + 0.43 versus controls). The grade 3 group (3 pillars, n = 19) sat below grade 2, consistent with sampling noise at small n; the trend across the well-populated grades 0 to 2 was unambiguous.

**Figure 5.**
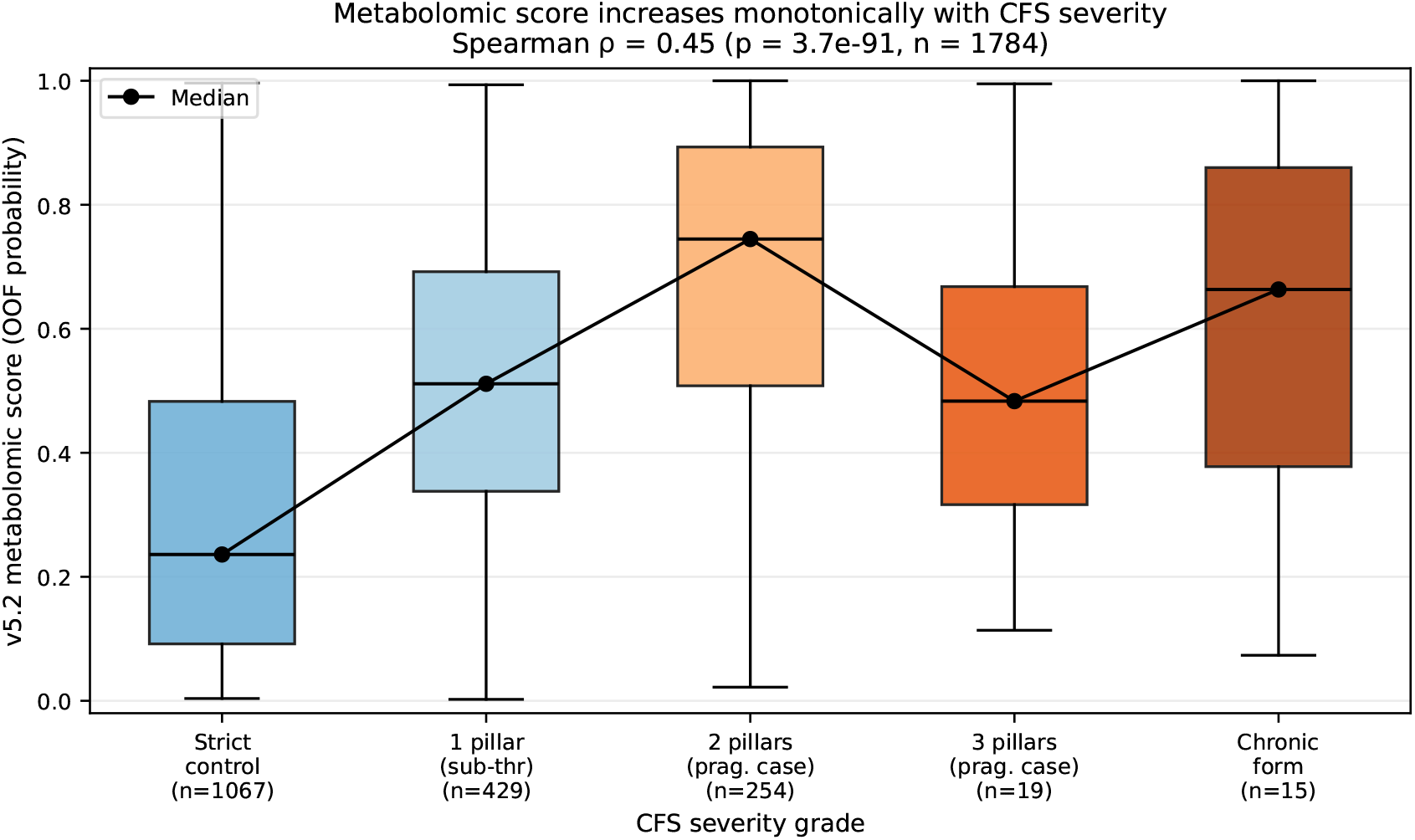
Severity stratification of the locked biology-first DBS score.

### 3.7 Hypothesis-free screening at scale captures exposome variance (Figure 6)

The 15 min univariate Wave B screen returned four features with cross-endpoint AUC ≥ 0.60 on all six endpoints, plus four additional candidates from the next ranks. Exact-mass database lookup of these eight leaders returned two Desmedipham annotations at slightly displaced retention times, one industrial NORMAN-SLE amine, two likely LC-MS artifacts (mobile-phase tributylamine and a probable charged-state or noise feature at m/z 144.545), one phytochemical annotation, one endogenous small-molecule candidate and one truncated oxidised phospholipid (1-palmitoyl-2-(5-hydroxyvaleroyl)-sn-glycero-3-phosphate; ChEBI confidence 0.74). None of the eight leaders had MS2 evidence in any of three independent MS2 directories. Inspection of precursor selection density in the relevant m/z windows indicated that these precursors were not selected for fragmentation. The majority of top Wave B leaders therefore appear to reflect exposome or technical variance rather than confirmed endogenous metabolism; the oxidised phospholipid remains a plausible but unconfirmed lead pending targeted MS2 or standard confirmation.

**Figure 6.**
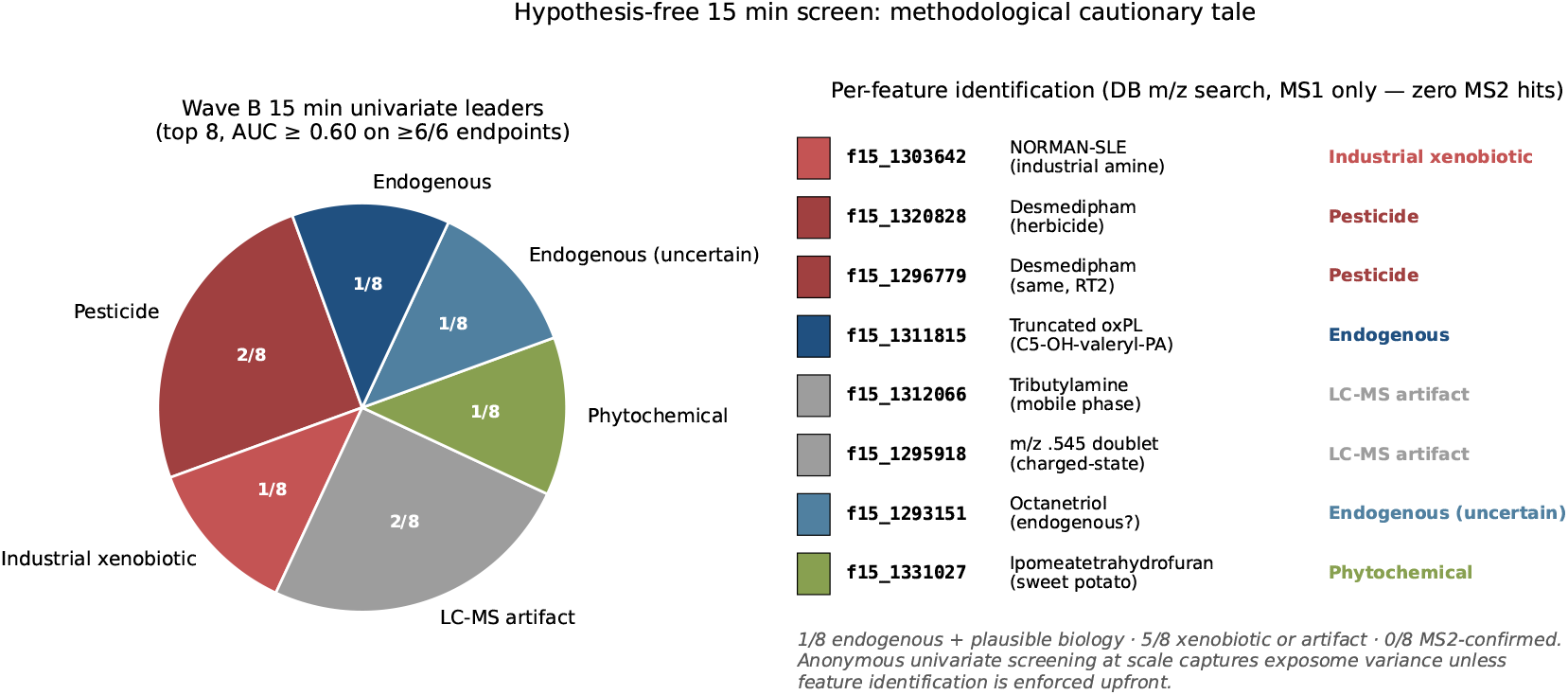
Identification review of hypothesis-free 15 min Wave B leaders.

### 3.8 Sensitivity analyses and anti-leak questionnaire baselines

The headline biology-first AUC reproduced within rounding on the full eligible set (Panel A AUC 0.805, 226 cases / 914 controls; bootstrap 95 % CI 0.766 to 0.839). On the covariate-complete QC subset (138 cases / 427 controls), DBS-only AUC was 0.762 (CI 0.709 to 0.811); demographics alone gave 0.636 (CI 0.584 to 0.687); DBS plus four covariates reached 0.781 (CI 0.730 to 0.831). The metabolomic lift over demographics on the same participants was therefore approximately 0.13 to 0.15 AUC units, and survived covariate adjustment (Table 1).

| Panel | n | n_pos | n_ctrl | Features | AUC | 95 % CI | |_|_|_|_|_|_|_| | A. DBS-only, full eligible | 1,140 | 226 | 914 | 88 | 0.805 | 0.766 to 0.839 | | B. DBS-only, QC subset | 565 | 138 | 427 | 88 | 0.762 | 0.709 to 0.811 | | C. DBS + covariates, QC subset | 565 | 138 | 427 | 92 | 0.781 | 0.730 to 0.831 | | D. Covariates only, QC subset | 565 | 138 | 427 | 4 | 0.636 | 0.584 to 0.687 | | E. Anti-leak clean covariate baseline, QC subset | 565 | 138 | 427 | 5 | 0.630 | 0.578 to 0.680 | | F. Anti-leak declarative cross-form baseline, QC subset | 565 | 138 | 427 | 7 | 0.631 | 0.579 to 0.684 | | G. F + allow_sensitivity_only items, QC subset | 565 | 138 | 427 | 9 | 0.669 | 0.617 to 0.722 |

Table 1. Sensitivity analyses on the locked primary endpoint (`pragmatic_fatigue_pem`), all panels evaluated with the same model spec (`Simpleimputer(median)` then `RobustScaler` then `RidgeClassifier(class_weight=“balanced”, alpha=10.0)`; five-fold stratified out-of-fold cross-validation; AUC on `sigmoid(decision_function)`; bootstrap 95 % CIs from 1,000 resamples). The QC subset is the `covariate_complete_core_qc` set (age, sex, BMI in plausible ranges).

Anti-leak questionnaire baselines did not substitute for the DBS panel. The clean covariate baseline (Panel E) reached AUC 0.630, and adding the two declarative cross-form items that pass the anti-leak filter for this endpoint (Panel F) raised it to 0.631. Both remained materially below DBS-only on the same QC subset (Panel B AUC 0.762), leaving a metabolomic-specific lift of approximately 0.131 AUC over the strongest anti-leak questionnaire baseline.

Subgroup DBS-only out-of-fold AUC was stable: 0.797 in women and 0.800 in men; 0.824 in age < 50 and 0.763 in age ≥ 50; 0.708, 0.855 and 0.801 across BMI strata < 25, 25 to 30 and ≥ 30. The overlapping bootstrap intervals indicated that no single demographic subgroup was carrying the signal alone.

The 1:4 nearest-neighbour matched-control analysis on age (± 3 years), sex (exact) and BMI (± 2 kg /m^2) retained 152 cases and 289 controls; DBS-only out-of-fold AUC on the matched set was 0.715 (95 % CI 0.665 to 0.767). The drop from 0.805 on the full eligible set is consistent with the loss of statistical power in a smaller, demographically restricted comparison rather than with covariate-driven inflation. Per-panel summaries, subgroup AUCs and the matched-control output are provided in `v4_sensitivity_panels_summary.csv`, `v4_sensitivity_subgroups.csv` and `v4_sensitivity_matched.csv`.

### 3.9 Longitudinal feasibility through dense self-sampling

One participant (BT1018) contributed thousands of self-collected fingerstick DBS samples over an extended observation window alongside continuous wearable-device streams (sleep, activity, heart rate variability) and daily body-weight measurements. The trajectory cannot provide population-level validation and is reported as supplementary hypothesis-generating material only (S8). It is included here in the main text as an operational result rather than as an inferential one: it demonstrates that dense longitudinal self-sampling at this granularity is feasible in an outpatient research setting and that the metabolomic, wearable and body-weight streams can be aligned on a shared per-day index. This case provides a framework for future within-person analyses of metabolic variation against sleep, activity, recovery and body-weight dynamics, and for tracking PEM-like episodes within an individual over time. The associated biological interpretation remains participant-specific and event-sensitive and is not generalised to the cohort.

## 4 Discussion

This study provides three observations about DBS metabolomic biology in chronic fatigue and one methodological observation about untargeted discovery in large community cohorts.

### The pathway-level signal is reproducible in DBS and concentrated on a PEM-like phenotype

A biology-first DBS panel of 22 metabolites represented by 88 engineered descriptors reached out-of-fold AUC 0.81 on a 226-case pragmatic PEM-like endpoint, showed pathway-level convergence with axes reported by Naviaux 2016 [8], Germain 2017 to 2022 [10, 11, 13] and Che 2022 [15], and scaled with severity (Spearman rho = 0.45, p < 10^−90^). The full cohort is approximately nine-fold larger than the largest prior plasma or serum metabolomics cohort in this literature (Che 2022, n = 197 [15]) and more than 20-fold larger than the original Naviaux discovery cohort [8]. This scale shifts the primary question from whether a pathway-level signal can be detected to how much of the PEM-associated signal is captured by a compact, literature-curated panel in routine DBS metabolomics. In this dataset, extending the curated panel with additional pathway-targeted features did not improve performance on the pragmatic PEM-like endpoint, suggesting that the core 22-metabolite panel captures most of the available signal accessible to this reverse-phase DBS platform for that endpoint.

### The signal is endpoint-specific to PEM-like symptom structure

Pragmatic PEM-like status (AUC 0.81), comorbid fatigue (0.73) and the DSQ-derived PEM-like construct (0.60 on the un-filtered set, 0.778 in the non-comorbid signature-decomposition follow-up of Section 3.2) encode either PEM logic or metabolic context, and these were the endpoints with the strongest metabolic signal. The pragmatic endpoint is not a validated diagnostic instrument; it is a higher-powered operationalisation of IOM-like PEM pillars. The DSQ-derived construct provides stronger questionnaire construct anchoring but has substantially fewer cases in this cohort. The gap between the un-filtered and non-comorbid DSQ-PEM estimates indicates that approximately half of the DSQ-PEM-positive participants carry at least one self-declared major-metabolic-history tag, so part of the headline 0.81 versus the un-filtered 0.60 reflects exclusion versus inclusion of declared comorbidity rather than an intrinsic metabolic ceiling on DSQ-PEM. Vithin this non-comorbid construct, an optimised operationalisation combining panel-self normalisation, the non-comorbid restriction and demographic covariates reached AUC 0.71 (Section 3.2), and an exploratory age-stratified analysis indicated that per-signature decomposition recovers separation in adults aged 40 and over (AUC 0.72) that a single pooled panel misses (AUC 0.62), with the dominant pathway block shifting with age; because the youngest and oldest strata each rest on only seven cases, these age-stratified phenotypes are hypothesis-generating and require independent confirmation. The lower un-filtered DSQ-derived AUC nonetheless constrains the diagnostic interpretation of the pragmatic 0.81 estimate. These data support a PEM-like self-report phenotype, rather than fatigue in general, as the phenotype most strongly captured by this metabolomic panel. This is consistent with cellular-bioenergetic [25, 26] and post-exercise [13, 20] literature in which exertional intolerance is the load-bearing physiological event. For downstream biomarker cohorts, case definition should therefore be treated as a primary design variable, not a secondary annotation.

### The signal is technically concordant across orthogonal LC gradients

Running the same DBS extracts through 5 min and 15 min reverse-phase gradients and computing per-metabolite univariate AUC independently in each gradient yielded Pearson r = 0.62 on signed effects (p = 0.002, approximate 95 % CI 0.27 to 0.83, 68 % sign agreement, n = 22). The strongest concordant signals align with tryptophan-kynurenine [16], redox-thiol [12] and amino-acid [4, 9] axes reported in prior ME/CFS metabolomics studies. For DBS, however, redox-thiol interpretation must account for cellular blood components, particularly erythrocyte-rich glutathione. This cross-gradient result is technical concordance rather than independent population validation, but it materially strengthens the evidence that the panel is not a single-run chromatographic artifact.

### Hypothesis-free screening at scale captures exposome variance

The 15 min univariate screen tested 5,625 features against six endpoints with cross-stability filtering and returned four leaders with AUC ≥ 0.60 on all six endpoints. Exact-mass annotation of the top eight candidates mapped most leaders to pesticide, industrial-amine or LC-MS artifact candidates; only one, a truncated oxidised phospholipid, was biologically plausible, and none had MS2 evidence in any of three independent MS2 directories. All eight therefore remain at Schymanski Level 4 to 5 [24]. At cohort sizes above 1,000 participants, untargeted univariate screening will reliably surface features that distinguish cases from controls. In community cohorts, some of those features may reflect exposome or technical variance rather than endogenous disease biology. Discovery pipelines at this scale should therefore require molecular identification before mechanistic interpretation.

### The signal is robust to demographic, behavioural and declarative-questionnaire confounding

On the covariate-complete subset (n = 565), age, sex, BMI and an additive medication burden, jointly or as a stand-alone demographic ceiling, did not reproduce the DBS panel: demographics-only reached AUC 0.64, DBS-only reached 0.76 and DBS plus covariates reached 0.78, leaving a metabolomic lift over demographics on the order of 0.13 to 0.15 AUC. The anti-leak declarative cross-form baseline, built from the questionnaire items that pass the 2026-05-26 anti-leak matrix for the pragmatic PEM-like endpoint, reached AUC 0.63, leaving a metabolomic-specific lift of approximately 0.13 AUC over the strongest allowed questionnaire baseline. Subgroup DBS-only AUC was stable across sex, age and BMI strata, and a 1:4 matched-control analysis on age, sex and BMI gave AUC 0.72. Taken together, these sensitivity analyses indicate that the DBS signal is not explainable by demographic confounding, by the declarative cross-form questionnaire variables that pass the anti-leak filter, or by a single demographic subgroup. The conservative matched-control estimate (AUC 0.72) is consistent with reduced power on a smaller demographically restricted set and with the cross-gradient and severity-trend results above, rather than with covariate-driven inflation of the headline AUC.

### At-home-compatible sampling is operationally relevant for the affected population

Fingerstick DBS does not require phlebotomy and is compatible with home self-collection. For fatigue and PEM-like syndromes, where repeated clinic attendance can itself trigger or aggravate symptoms, this format reduces the access barrier to repeated biological measurement and supports within-person longitudinal monitoring (illustrated for one participant in Section 3.9). The clinical-translation implications of an at-home metabolomic layer, including the independent-validation steps required before any decision-support deployment, are detailed in Section 5.

## 5 Clinical translation and validation roadmap

For clinical translation, the DBS score described here should be considered a candidate biological decision-support layer pending independent validation, not a stand-alone diagnostic test. The intended use case is to complement clinician assessment and questionnaire-based phenotyping with an at-home-compatible biological measurement that can be repeated longitudinally. Future studies should evaluate whether the locked score improves clinician assessment beyond symptom questionnaires, differentiates PEM-like fatigue from other fatigue etiologies, and tracks within-person changes in function, recovery or post-exertional worsening over time.

The next step is independent validation in a clinically adjudicated cohort that includes confirmed ME/CFS cases (Canadian Consensus or IOM 2015 criteria with clinical adjudication), fatigue controls without ME/CFS and non-fatigued controls. The locked DBS score should be tested without retraining, with pre-specified evaluation of calibration, decision thresholds, test-retest repeatability on repeated self-collected DBS, and paired DBS-versus-plasma or DBS-versus-serum comparability on a subset to clarify matrix-effect interpretation. At-home and clinic-collected samples on the same participants would also clarify the operational equivalence of self-collected versus phlebotomy-derived DBS for this score. Adjudicated subgroup analyses should be pre-specified for comorbid presentations including POTS, MCAS, fibromyalgia and hypothyroidism, which were not isolated in the present cohort (Limitation 7). Until these steps are completed, the present results should be communicated as exploratory platform evidence supporting at-home DBS metabolomics as a biologically grounded substrate for future clinically adjudicated validation and decision-support development, rather than as evidence for a diagnostic test, screening tool or objective provoked-PEM biomarker.

## Data Availability

Raw mass-spectrometry data and individual participant-level metadata cannot be released publicly because of participant privacy and consent constraints attached to a commercial research program. They are available from the corresponding author on reasonable request, subject to a data-use agreement.

## 6 Limitations

1. This is a single-cohort study without an independent external cohort. The cross-gradient analysis provides technical concordance, not independent population validation.
2. The analysed matrix is DBS, not venous plasma. DBS contains cellular and plasma-derived components, so individual metabolite comparisons with plasma or serum ME/CFS studies should be interpreted at pathway level rather than as direct concentration equivalence. This is especially important for redox-thiol features such as glutathione, which may be strongly influenced by erythrocyte content.
3. Endpoints were derived from structured questionnaires. The pragmatic PEM-like endpoint provides power but is not a validated diagnostic instrument; the DSQ-derived construct provides stronger questionnaire anchoring but had only 57 cases. Provoked PEM was not available here. Symptom-day alignment between blood collection and questionnaire response may also vary across participants.
4. Questionnaire analyses require explicit anti-leak separation. The 2026-05-26 anti-leak matrix resolves the item-classification step, and the clean covariate-only and declarative cross-form baselines have been computed and reported in Section 3.8. They remain self-report baselines: they assess concordance with allowed declarative items, not independent clinical validation.
5. Sensitivity to demographic and treatment confounding has been assessed (Section 3.8): age, sex, BMI and a medication-burden covariate were adjusted on the covariate-complete subset, subgroup AUCs were computed by sex, age and BMI, and a matched-control AUC was reported. Menopausal status was not separately modelled, and unmeasured behavioural or treatment confounders cannot be fully excluded.
6. Participants were enrolled from a BioTwin community cohort of individuals who provided samples and questionnaires through a commercial research program. The cohort is therefore not population-representative, and generalisability to clinically ascertained ME/CFS cohorts or asymptomatic screening populations is limited.
7. Comorbid presentations of ME/CFS, including POTS, MCAS, fibromyalgia and hypothyroidism, were not isolated in this analysis. Some of the metabolic signal attributed to PEM may co-vary with these comorbidities.
8. No stratification by illness duration, treatment exposure or symptom subtype was performed.
9. The chronic-fatigue-form endpoint (n = 12 cases) is too small for reliable multivariate inference; AUCs on this endpoint should be treated as exploratory.
10. DDA-MS2 coverage was incomplete for several leads. In particular, the truncated oxidised phospholipid lead (m/z 509.288, RT 10.88 min) had zero DDA scans selected in its m/z window across the available 15 min batches. Targeted MS2 re-acquisition or standard confirmation is required before this candidate can be promoted.
11. The hypothesis-free Wave B leaders are MS1-annotated only and should not be interpreted as confirmed metabolites.
12. The BT1018 dense-sampling case study is retained as supplementary hypothesis-generating material only. It is participant-specific, event-sensitive and not a population-level validation.
13. The DSQ-PEM signature-decomposition follow-up reported in Section 3.2 was run on a smaller cohort (n = 259; 29 cases / 230 controls) obtained by excluding participants flagged by `has_major_meta`. This flag is constructed from substring matches on a free-text onboarding history form covering cancer, autoimmune, cardiac, diabetic, ulcer, depressive, neurological and joint history. In the full BioTwin base of 1,784 participants, 22.4 % carry at least one such tag, and within the `label_screened_control` pool the same filter excludes essentially zero participants because the screened-control construction already enforces absence of these histories. The filter therefore removes approximately half of the DSQ-PEM-positive participants while leaving the control pool unchanged, which shifts the comparison toward a non-comorbid case versus non-comorbid control contrast. The flag reflects self-reported history at onboarding rather than active clinical diagnosis, and the analysis should be interpreted as isolating a non-comorbid PEM-like phenotype rather than as a clinically adjudicated comorbidity exclusion.
14. The DSQ-PEM signature-decomposition AUC (0.778, Section 3.2) was estimated at the participant level, with descriptors aggregated across all available cards per participant. The corresponding per-card evaluation using five-fold GroupKFold on participant identifier reached AUC 0.64. This approximately 0.14 AUC gap indicates that the score is supported by aggregation across repeated sampling occasions and should not be deployed as a single-card readout in user-facing applications.
15. The `n_nonmissing_cards` participant-level descriptor adds approximately 0.02 AUC to the late-fusion DSQ-PEM follow-up overall, but its univariate AUC on the age < 40 sub-stratum reaches 0.86. Sampling cadence may therefore covary with clinical state in younger participants and should be tracked separately from biochemical descriptors when reporting age-stratified results.
16. The DSQ-PEM operationalisation-improvement and age-stratification analyses (Section 3.2) are post-hoc and exploratory. The improved “C6” specification and the four-signature late fusion were developed and reported on the same reduced non-comorbid cohort (n = 259; 29 cases) and were not validated out-of-cohort; their gains over the locked specification should be read as optimistic upper bounds pending independent replication. The age-stratified estimates for the under-40 and 60-and-over strata rest on seven cases each, with wide confidence intervals, and must not be interpreted as established age-specific phenotypes.

## 7 Data and code availability

- Raw mass-spectrometry data and individual participant-level metadata cannot be released publicly because of participant privacy and consent constraints attached to a commercial research program. They are available from the corresponding author on reasonable request, subject to a data-use agreement.
- Aggregated outputs supporting this paper (the locked biology-first panel definition, the per-panel sensitivity summary, the per-metabolite cross-gradient match table and the pathway-level AUC matrix) are available from the corresponding author on reasonable request.
- Analysis code, including the 5 min and 15 min coverage filters, the feature-by-batch robust z-scoring routines, the Ridge out-of-fold evaluation pipeline, the bootstrap stability framework and the v4 sensitivity-analysis script (`run_v4_sensitivity_and_crossform.py`), is available from the corresponding author on reasonable request.

## 8 Funding and competing interests

This work was funded entirely by BioTwin Inc. through internal research and development. No external government grants or agency funding were received for this study.

All authors are employees and shareholders of BioTwin Inc. N.A. serves as Chief Scientific Officer, P.H. serves as Chief Technology Officer, and L.-P.N. serves as Chief Executive Officer and Founder. BioTwin Inc. develops metabolomic profiling and digital-twin technologies that may have commercial relevance to the work described here. The authors declare that the scientific conclusions presented in this manuscript are not influenced by these financial interests. No independent external validation of the results has been performed by parties external to BioTwin Inc.

## Figure list

- Figure 1. Cohort, pipeline and endpoint cardinalities. `fig1_cohort.pdf/png`
- Figure 2. v5.2 OOF AUC by endpoint. `fig2_endpoint_specificity.pdf/png`
- Figure 3. Pathway-level forest plot and heatmap (Wave A). `fig3_pathway_forest.pdf/png`
- Figure 4. Cross-gradient 5 min vs 15 min per-metabolite AUC scatter. `fig4_cross_val_5min_15min.pdf`
- **Figure 4b:** Pathway convergence with prior metabolomics cohorts. `fig4b_pathway_convergence.pdf/pn`
- Figure 5. Severity stratification of v5.2 score. `fig5_severity_stratification.pdf/png`
- Figure 6. Hypothesis-free Wave B 15 min leaders identification breakdown. `fig6_wave_b_exposome.pdf/`

## Supplementary material

- **S1:** `cfs_v5_2_literature_expansion_user_features.csv` (88 v5.2 user-level features x 1,784 btids; restricted access).
- **S2:** `cfs_v8_wave_a_feature_block_metrics.csv` (28 pathway blocks x 6 endpoints; AUC, AP, n_features, reason).
- **S3:** `fig3_pathway_forest_data.csv` (pathway-level AUC matrix).
- **S4:** `cross_val_5min_15min_best_match.csv` (22-metabolite cross-gradient table).
- **S5:** `severity_stratification_summary.csv` (severity grade x score statistics).
- **S6:** `cfs_v8_wave_b_15min_leaders_top3_idents.csv` (Wave B leaders identification details, MS1 only).
- **S7:** `phase1_outputs/phase1_a2_questionnaire_items_VALiDATED_2026-05-26.csv` (anti-leak questionnaire matrix).
- **S8:** BT1018 dense-sampling trajectory outputs, supplementary hypothesis-generating material only.
- **S9:** v4 sensitivity outputs (`v4_sensitivity_panels_summary.csv`, `v4_sensitivity_subgroups.csv`, `v4_sensitivity_matched.csv`).
- **S10:** DSQ-PEM signature-decomposition follow-up (Section 3.2): per-signature feature lists (S1, S2, S4, S5), per-seed late-fusion AUC, leave-one-out BT1018 sensitivity, per-card GroupKFold result, and per-stratum `n_nonmissing_cards` AUC. Source scripts: `dsq_improve_two_signatures.py`, `dsq_improve_multi_signatures.py`, `dsq_audit_outlier_and_pers dsq_audit_leakage_test.py`.
- **S11:** Biology-first metabolite panel annotation confidence and literature convergence (`cfs_v4_3_panel_annotation_confidence.csv`). One row per biology-first metabolite (n = 22) with columns: Metabolite, Pathway, m/z (theoretical), Retention time (5 min and 15 min, min), Gradient(s) used, Descriptor set, Annotation confidence (Schymanski / MSI level [24]), MS2 evidence (yes/no, source MS2 directory), Direction in DBS (cases versus controls), Direction in prior ME/CFS literature, Primary references, and DBS matrix caveat.
- **S12:** DSQ-PEM operationalisation-improvement and age-stratification results (Section 3.2). Per-variant out-of-fold AUCs for the normalisation, cohort-restriction and covariate levers and the stacked “C6” specification (`dsq_improve_combined_results.csv`, `dsq_improve_all_vari` and per-stratum per-signature late-fusion AUCs by age (`dsq_improve_age_stratified_results.csv`). Source scripts: `dsq_improve_harness.py`, `dsq_improve_combined.py`, `dsq_improve_multi_signatures dsq_improve_age_stratified.py`.

## Decisions still pending before submission

1. Confirm author order, corresponding author and CRediT roles before submission.
2. Retain the pragmatic PEM-like status and DSQ-derived construct as separate endpoint constructs: one powered, one questionnaire-anchored.
3. Retain the truncated oxPL lead (`f15_1311815`) as discussion-only pending targeted MS2 or standard *confirmation*.
4. BT1018 N = 1 case study is referenced in Section 3.9 as a longitudinal-feasibility result with dense-sampling outputs retained in supplement S8; promote no inferential claim to the main text.
5. Target journal: *Metabolites* as primary; *Journal of Proteo e Researc* or *Frontiers in Molecular Biosciences* as alternates within the same applied-metabolomics class.

## Notes

### Author Declarations

Ethics committee/IRB of Canadian SHIELD Ethics Review Board gave ethical approval for this work (REB Tracking Number: 2023-11-003; OHRP Registration IORG0003491; FDA Registration IRB00004157; initial approval granted December 15, 2020; continuing review approval granted April 30, 2025, valid through April 29, 2026). All participants provided written informed consent prior to enrollment.

## References

[1] Institute of Medicine. Beyond Myalgic Encephalomyelitis/Chronic Fatigue Syndrome: Re-defining an Illness. Washington, DC: The National Academies Press, 2015. ISBN: 978-0-309-31689-7. DOI: 10.17226/19012.

[2] Leonard A. Jason and Madison Sunnquist. “The Development of the DePaul Symptom Questionnaire: Original, Expanded, Brief, and Pediatric Versions”. In: Frontiers in Pediatrics 6 (2018), p. 330. DOI: 10.3389/fped.2018.00330.

[3] Kjetil G. Brurberg, Marita S. FØnhus, Lillebeth Larun, Signe Flottorp, and Kirsti Mal-terud. “Case definitions for chronic fatigue syndrome/myalgic encephalomyelitis (CFS/ME): a systematic review”. In: BMJ Open 4.2 (2014), e003973. DOI: 10.1136/bmjopen-2013-003973.

[4] Teilah K. Huth, Natalie Eaton-Fitch, Donald Staines, and Sonya Marshall-Gradisnik. “A systematic review of metabolomic dysregulation in Chronic Fatigue Syndrome/Myalgic Encephalomyelitis/Systemic Exertion Intolerance Disease (CFS/ME/SEID)”. In: Journal of Translational Medicine 18 (2020), p. 198. DOI: 10.1186/s12967-020-02356-2.

[5] Hirohiko Kuratsune, Kouzi Yamaguti, Masakazu Takahashi, Hidehiko Misaki, Setsuko Tagawa, and Tetsuo Kitani. “Acylcarnitine deficiency in chronic fatigue syndrome”. In: Clinical Infectious Diseases 18 Suppl 1 (1994), S62–S67. DOI: 10.1093/clinids/18.supplement_1.s62.

[6] Stephanie E. Reuter and Allan M. Evans. “Long-chain acylcarnitine deficiency in patients with chronic fatigue syndrome. Potential involvement of altered carnitine palmitoyltransferase-I activity”. In: Journal of Internal Medicine 270.1 (2011), pp. 76–84. DOI: 10.1111/j.1365-2796.2010.02341.x.

[7] Christopher W. Armstrong, Neil R. McGregor, Donald P. Lewis, Henry L. Butt, and Paul R. Gooley. “Metabolic profiling reveals anomalous energy metabolism and oxidative stress pathways in chronic fatigue syndrome patients”. In: Metabolomics 11.6 (2015), pp. 1626–1639. doi: 10.1007/s11306-015-0816-5.

[8] Robert K. Naviaux, Jane C. Naviaux, Kefeng Li, A. Taylor Bright, William A. Alaynick, Lin Wang, Asha Baxter, Neil Nathan, Wayne Anderson, and Eric Gordon. “Metabolic features of chronic fatigue syndrome”. In: Proceedings of the National Academy of Sciences 113.37 (2016), E5472–E5480. Doi: 10.1073/pnas.1607571113.

[9] Emi Yamano, Masahiro Sugimoto, Akiyoshi Hirayama, Satoshi Kume, Masanori Yamato, Guanghua Jin, Seiki Tajima, Nobuhito Goda, Kentaro Iwai, Sanae Fukuda, Kouzi Yamaguti, Hirohiko Kuratsune, Tomoyoshi Soga, Yasuyoshi Watanabe, and Yosky Kataoka. “Index markers of chronic fatigue syndrome with dysfunction of TCA and urea cycles”. In: Scienti : c Reports 6 (2016), p. 34990. doi: 10.1038/srep34990.

[10] Arnaud Germain, David Ruppert, Susan M. Levine, and Maureen R. Hanson. “Metabolic profiling of a myalgic encephalomyelitis/chronic fatigue syndrome discovery cohort reveals disturbances in fatty acid and lipid metabolism”. In: Molecular BioSystems 13.2 (2017), pp. 371–379. doi: 10.1039/c6mb00600k.

[11] Arnaud Germain, Dinesh K. Barupal, Susan M. Levine, and Maureen R. Hanson. “Comprehensive Circulatory Metabolomics in ME/CFS Reveals Disrupted Metabolism of Acyl Lipids and Steroids”. In: Metabolites 10.1 (2020), p. 34. DOI: 10.3390/metabo10010034.

[12] Arnaud Germain, David Ruppert, Susan M. Levine, and Maureen R. Hanson. “Prospective Biomarkers from Plasma Metabolomics of Myalgic Encephalomyelitis/Chronic Fatigue Syndrome Implicate Redox Imbalance in Disease Symptomatology”. In: Metabolites 8.4 (2018), p. 90. DOI: 10.3390/metabo8040090.

[13] Arnaud Germain, Ludovic Giloteaux, Ryan A. Moriarty, Daniel R. Williams, Susan M. Levine, and Maureen R. Hanson. “Plasma metabolomics reveals disrupted response and recovery following maximal exercise in myalgic encephalomyelitis/chronic fatigue syndrome”. In: JCI Insight 7.9 (2022), e157621. DOI: 10.1172/jci.insight.157621.

[14] Dorottya Nagy-Szakal, Brent L. Williams, Nischay Mishra, Xiaoyu Che, Bohyun Lee, Lucinda Bateman, Nancy G. Klimas, Anthony L. Komaroff, Susan Levine, Jose G. Montoya, Daniel L. Peterson, Devi Ramanan, Komal Jain, Meredith L. Eddy, Mady Hornig, and W. Ian Lipkin. “Insights into myalgic encephalomyelitis/chronic fatigue syndrome phenotypes through comprehensive metabolomics”. In: Scientific Reports 8 (2018), p. 10056. DOI: 10.1038/s41598-018-28477-9.

[15] Xiaoyu Che, Christopher R. Brydges, Yuanzhi Yu, Adam Price, Shreyas Joshi, Ayan Roy, Bohyun Lee, Dinesh K. Barupal, Aaron Cheng, Dana M. Palmer, Susan Levine, Daniel L. Peterson, Suzanne D. Vernon, Lucinda Bateman, Mady Hornig, Jose G. Montoya, Anthony L. Komaroff, Oliver Fiehn, and W. Ian Lipkin. “Metabolomic Evidence for Peroxisomal Dysfunction in Myalgic Encephalomyelitis/Chronic Fatigue Syndrome”. In: International Journal of Molecular Sciences 23.14 (2022), p. 7906. DOI: 10.3390/ijms23147906.

[16] Bahar Kavyani, Brett A. Lidbury, Richard Schloeffel, Paul R. Fisher, Daniel Missailidis, Sarah J. Annesley, Mona Dehhaghi, Benjamin Heng, and Gilles J. Guillemin. “Could the kynurenine pathway be the key missing piece of Myalgic Encephalomyelitis/Chronic Fatigue Syndrome (ME/CFS) complex puzzle?” In: Cellular and Molecular Life Sciences 79.8 (2022), p. 412. DOI: 10.1007/s00018-022-04380-5.

[17] Hussein K. Al-Hakeim, Tabarek H. Al-Naqeeb, Abbas F. Almulla, and Michael Maes. “Tryptophan catabolites, inflammation, and insulin resistance as determinants of chronic fatigue syndrome and affective symptoms in long COVID”. In: Frontiers in Molecular Neuroscience 16 (2023), p. 1194769. DOI: 10.3389/fnmol.2023.1194769.

[18] Yamiló Lõpez-Hernández, Joel Monárrez-Espino, David Alejandro García Lõpez, Jiamin Zheng, Juan Carlos Borrego, Claudia Torres-Calzada, Josó Pedro Elizalde-Díaz, Rupasri Mandal, Mark Berjanskii, Eduardo Martínez-Martínez, Jesús Adrián Lõpez, and David S. Wishart. “The plasma metabolome of long COVID patients two years after infection”. In: Scientific Reports 13 (2023), p. 12420. DOI: 10.1038/s41598-023-39049-x.

[19] Anthony L. Komaroff and W. Ian Lipkin. “ME/CFS and Long COVID share similar symptoms and biological abnormalities: road map to the literature”. In: Frontiers in Medicine 10 (2023), p. 1187163. DOI: 10.3389/fmed.2023.1187163.

[20] Brent Appelman, Braeden T. Charlton, Richie P. Goulding, Tom J. Kerkhoff, Ellen A. Breedveld, Wendy Noort, Carla Offringa, Frank W. Bloemers, Michel van Weeghel, Bauke V. Schomakers, Pedro Coelho, Jelle J. Posthuma, Eleonora Aronica, W. Joost Wiersinga, Michòle van Vugt, and Rob C. I. Wüst. “Muscle abnormalities worsen after post-exertional malaise in long COVID”. In: Nature Communications 15 (2024), p. 17. DOI: 10.1038/s41467-023-44432-3.

[21] World Medical Association. “World Medical Association Declaration of Helsinki: ethical principles for medical research involving human subjects”. In: JAMA 310.20 (2013), pp. 2191–2194. DOI: 10.1001/jama.2013.281053.

[22] Hannes L. Röst, Timo Sachsenberg, Stephan Aiche, Chris Bielow, Hendrik Weisser, Fabian Aicheler, Sandro Andreotti, Hans-Christian Ehrlich, Petra Gutenbrunner, Erhan Kenar, Xiao Liang, Sven Nahnsen, Lars Nilse, Julianus Pfeuffer, George Rosenberger, Marc Rurik, Uwe Schmitt, Johannes Veit, Mathias Walzer, David Wojnar, Witold E. Wolski, Oliver Schilling, Jyoti S. Choudhary, Lars Malmström, Ruedi Aebersold, Knut Reinert, and Oliver Kohlbacher. “OpenMS: a flexible open-source software platform for mass spectrometry data analysis”. In: Nature Methods 13.9 (2016), pp. 741–748. DOI: 10.1038/nmeth.3959.

[23] Fabian Pedregosa, Gaël Varoquaux, Alexandre Gramfort, Vincent Michel, Bertrand Thirion, Olivier Grisel, Mathieu Blondel, Peter Prettenhofer, Ron Weiss, Vincent Dubourg, Jake Vanderplas, Alexandre Passos, David Cournapeau, Matthieu Brucher, Matthieu Perrot, and Édouard Duchesnay. “Scikit-learn: Machine Learning in Python”. In: Journal of Machine Learning Research 12 (2011), pp. 2825–2830.

[24] Emma L. Schymanski, Junho Jeon, Rebekka Gulde, Kathrin Fenner, Matthias Ruff, Heinz P. Singer, and Juliane Hollender. “Identifying small molecules via high resolution mass spectrometry: communicating confidence”. In: Environmental Science S Technology 48.4 (2014), pp. 2097–2098. DOI: 10.1021/es5002105.

[25] Cara Tomas, Audrey Brown, Victoria Strassheim, Joanna L. Elson, Julia Newton, and Philip Manning. “Cellular bioenergetics is impaired in patients with chronic fatigue syndrome”. In: PLoS ONE 12.10 (2017), e0186802. DOI: 10.1371/journal.pone.0186802.

[26] Norman E. Booth, Sarah Myhill, and John McLaren-Howard. “Mitochondrial dysfunction and the pathophysiology of Myalgic Encephalomyelitis/Chronic Fatigue Syndrome (ME/CFS)”. In: International Journal of Clinical and Ezperimental Medicine 5.3 (2012), pp. 208–220.

[27] Robert K. Naviaux et al. “Correction for Naviaux et al., Metabolic features of chronic fatigue syndrome”. In: Proceedings of the National Academy of Sciences 114.18 (2017), E3749. DOI: 10.1073/pnas.1615143113.

